# “*Just a knife wound this week, nothing too painful”:* an ethnographic exploration of how homeless clients attending an urban primary care and addiction service view their own health and healthcare

**DOI:** 10.1101/2024.02.19.24302966

**Authors:** Carolyn Ingram, Conor Buggy, Isobel MacNamara, Carla Perrotta

**Affiliations:** Public Health, School of Public Health, Physiotherapy, and Sports Science, University College Dublin, Dublin, Ireland; School of Medicine, University College Dublin, Dublin, Ireland

## Abstract

This study used an ethnographic approach grounded in a social constructivist research paradigm to explore the perspectives of people in homelessness attending a primary care and addiction service on their priority health and healthcare needs. Participant observations and informal interviews were conducted with homeless clients attending the service for three hours every Monday morning between October 2022 and April 2023. Field note data from active participant observation and informal conversations were collected, anonymised, and analysed using inductive thematic analysis in accordance with the Declaration of Helsinki and the researchers’ institutional Research Ethics Committee. Three main themes emerged from the analysis: self-identified priorities, satisfaction with services, and migrant health. Clients’ priority concerns relate to their mental health, maintaining ties with children and families, navigating complex romantic relationships, finding meaningful activities, and feeling better physically. These challenges differ from those of the general population in terms of their severity observed both prior to and during experiences of homelessness, coupled with disproportionately high levels of loss, fear, injury, pain, disability, fatigue, and isolation. In terms of services, clients are satisfied with their ability to access primary care and harm reduction in a social environment where positive exchanges with friends and providers take place. Conversely, barriers to accessing mental health and addiction services persist including the internalised belief that one is beyond help, lack of access to information on available services and their entry requirements, and lingering stigma within a health system that treats addiction as separate to health. Moving forward, health practitioners may consider holding more regular and open conversations with homeless clients about the care they are receiving, its rationale, and whether or not changes are desired that can be safely made. The health needs of migrants and asylum seekers entering homelessness in Ireland are urgent and should be prioritised in future research.

## Introduction

Rising homelessness levels across Europe have brought increased attention to the health consequences of precarious housing. People experiencing homelessness (PEH) are at higher risk of premature mortality, disability, and chronic conditions but face reduced access to health services [1, 2]. A health equity perspective – the belief that everyone has the right to a fair and just opportunity to attain their full potential for health – emphasizes that housing-related health disparities are unnecessary and avoidable. Rather than the result of biology, they result from social and economic processes that create / recreate differences in access to housing, health services, and in turn health[3]. Medical and public health practitioners acknowledge that advancing health equity requires not only addressing the downstream drivers of homelessness, but also improving equitable access to healthcare for PEH who experience myriad barriers to care due to complicated application processes or appointment systems, long queues, judgement from healthcare providers, competing priorities (e.g., finding food or a place to sleep), and internal barriers like fear, embarrassment, hopelessness, and poor self-esteem [4].

Ireland is unique in Europe in the severity of spikes in homelessness over the last decade [5]. Across the Dublin region (population 1.4 million) where most national homelessness is concentrated, approximately 6,700 adults and 3,000 children are currently residing in emergency accommodation [6]. An estimated 118 individuals are sleeping rough [7], and at least 6% of the city’s residents are experiencing hidden homelessness (i.e., sleeping cars, in squats, on the floors or sofas of family and friends, or in unsafe accommodation [8]. Drug and alcohol use is the primary driver of homelessness in Ireland and remains a leading cause of death within the homeless population [9]. In response, and to circumvent known barriers to care, a network of multi-disciplinary primary health care and addiction services provide specialised services for PEH across Dublin in partnership with the Health Services Executive Ireland (HSE) [10]. These services have drastically improved access to primary care and harm reduction in recent years and Ireland is seen to be a leader in Europe in its provision of innovative healthcare at the community level [10, 11].

Despite these improvements, in autumn 2022, multiple factors highlighted a need to identify priority health concerns amongst PEH who are accessing homeless health services in Ireland. First, despite increased access to primary care, evidence shows that disparities in mortality between homeless and housed populations continue to worsen [12]. Second, high rates of Covid-19 vaccine hesitancy amongst marginalised groups stressed the extent to which public health campaigns that fail to identify the priority needs and concerns of those whom they affect are counterproductive, especially amongst communities used to being let down by health and other government services [13]. Finally, the Covid-19 pandemic acted as a catalyst for change in the accessibility of harm reduction measures for homeless individuals who use drugs. Positive changes – namely increased access to methadone and naloxone – demonstrated the capacity for policymakers to remove barriers to accessing care in response to a strong public health argument [14]. Researchers thus saw an opportunity to provide further evidence on how and where to allocate health and social service resources effectively.

Community health needs assessments (CHNA) that include qualitative techniques are designed to help tailor health services to the needs of specific population groups [15]. The integration of voices from community members into these assessments enriches the CHNA process in several ways [16]. A social constructivist paradigm assumes the world is made up of multiple realities, local truths, and subjective experiences [17]. From this lens, interacting with community members provides otherwise unobservable data that may help bridge the divide between theory and practice [18]. Qualitative needs assessment approaches involving community members can help improve quality by identifying gaps or current problems in care and help focus initiatives on what is important to those for whom they are designed [16]. Labonte and Robinson[17] describe what may follow should health researchers and practitioners overlook the needs most important to a community, *“If we fail to start with what is close to people’s hearts by imposing our notions of health concerns over theirs, we risk several disabling effects. We may be irrelevant to the lives and conditions of many persons … We may further their experience of powerlessness by failing to listen and act upon concerns in their lives as they experience and name them, communicating to them that they are wrong and we are right.”* To date, there remains a gap in the evidence in terms of how PEH view their own health and healthcare in Ireland [11].

To ensure the integration of community voices into a larger, qualitative CHNA[16], this study used an ethnographic approach to explore the perspectives of people in homelessness attending an urban primary care and addiction service on their priority health and healthcare needs.

## Materials and methods

### Study Design

The lead researcher engaged in ethnographic fieldwork at a drop-in primary care and addiction services clinic serving primarily PEH. She initially approached the clinic founder to determine mutual interest in a research collaboration and establish a community partnership. The central question explored in this article (i.e., *What are PEH’s own views on their health and healthcare needs?*) and associated research methods were then developed by the research team and clinic staff. Ethnography was the chosen methodology grounded in a social constructivist research paradigm. Ethnographic methods allowed for a focus on holism[19]. Rather than predefining and restricting the areas the researcher considered relevant to the health of PEH, a broader approach allowed her to identify and understand client concerns she could not have expected. A social constructivist paradigm holds that multiple realities exist, each an intangible construction rooted in people’s experiences with everyday life, how they remember them, and make sense of them [20]. From this lens, the researcher could become a research instrument for generating a “consensus construction of reality” more informed and sophisticated than individual constructions[17, 20].

The ethnographic approach to fieldwork combined active participant observation and informal interviews – both casual and more in-depth – with clinic attendees. Participant observation was used to gain insight into naturally occurring data and events happening in the study setting and to identify how clients are accessing / using primary healthcare and addiction services. The researcher adopted the role of the ‘active participant observer’; her research was explicit rather than covert. This allowed her to become trusted by those attending and working at the clinic over time and keep returning patients apprised of her ongoing findings and activities [21]. Observations were enriched by informal conversations that helped the researcher strengthen trust, establish a rapport, and see situations from clients’ perspectives in a natural and inartificial way. Researchers argue that informal interviews build a less threatening environment, create an ease of communication, and allow for the engagement of vulnerable community members who may be less likely to participate in formal interviews or surveys [22, 23].

### Participants and setting

The target population for the ethnographic research was homeless individuals linked with primary healthcare and/or addiction services in Ireland. The sampled population was people experiencing (or who had experienced) homelessness attending the partner drop-in primary care and addiction services clinic at least one Monday between October 2022 and April 2023. For the purposes of this study, homelessness was defined as (1) having no accommodation available that can reasonably be stayed in or living in a hospital, county home, night shelter or other such institution, and (2) being unable to provide accommodation from one’s own resources[24]. The study site provides a walk-in general practitioner (GP) service on weekday mornings for patients who are experiencing homelessness or who are undocumented migrants. Key working services and needle exchange are provided on site, as are opiate substitution and – under a strict protocol – benzodiazepine detoxification and community alcohol detoxification. Patients who have a medical card (formal access to free health services through the HSE) are encouraged to seek care with their own GP.

Prior to commencing the formal observation period, the researcher visited the clinic to meet staff and familiarise herself with the setting. Clinic attendees were selected for informal interviews during the researcher’s subsequent visits using purposive, critical case sampling [25]. Clinic medical or operational staff identified and introduced the researcher to potential ‘critical cases’ who (1) had experienced/were experiencing homelessness and potential co-occurring addiction, (2) had an established rapport with centre staff and were deemed to be of sound mental capacity, (3) expressed a voluntariness to chat with the researcher without coercion, and (4) spoke English.

### Data Collection

Participant observations and informal interviews were conducted for three hours every Monday morning between October 2022 and April 2023. The day was selected based on clinic staff’s preferences; Mondays were deemed the least chaotic with more space for the researcher to chat with patients. The researcher chose not to record conversations during her visits as it could raise patients’ wariness toward the project [26]. Instead, she jotted down raw records in a notebook (during observations and in-depth conversations or after casual conversations). Within 24 hours of each visit, the researcher wrote up formal fieldnotes, with a detailed description of the visit, her personal reflections, emerging questions/analyses, and future actions. Data collection took place in the form of in-depth and casual conversations, and observations.

### In-Depth Conversations

After an introduction to a potential participant by a clinic staff member, the researcher presented herself as a PhD scholar interested in the participant’s concerns and needs regarding their healthcare. If the client agreed, they were invited to accompany the researcher into an open consultation room. Informed consent for informal interviews was obtained verbally; a recommended approach for non-recorded research with people who are/have been homeless and experience significant state regulation and stigma leading to mistrust of activities that require their signature [27]. Participants were informed in more detail that the research was looking at clients’ own concerns and needs regarding their healthcare, that any notes taken during and after the conversation would be fully anonymized, and that results would be submitted for scientific publication and shown to policymakers. Once informed verbal consent was obtained, the researcher led with a general question: *“Do you mind telling me a bit about how are you feeling today?”* If healthcare didn’t come up naturally in conversation, the researcher would ask, when appropriate, *“and what about your healthcare, do you feel like you could be better supported in any way?*” In-depth conversations lasted between 20 minutes to 1 hour.

### Casual Conversations

In some instances, the researcher would initiate a non-directive [26] conversation in the waiting room with the individual seated next to her, or vice versa. She learned simple techniques for determining an individual’s interest in conversing; things like saying ‘bless you’ when someone sneezed or offering to watch someone’s seat while they went to the toilet. If the person was quiet, the researcher would not try to engage them any further. If the person was interested in chatting, the researcher would follow the organic flow of conversation, documenting fully anonymised exchanges in her field notes after each visit. When appropriate, the researcher would formally introduce herself. Because she was not able to assess whether the individual was in a position to grasp and reflect on information about the study, she would not invite someone to participate in an in-depth conversation without a formal introduction from clinic staff. These conversations – by allowing the researcher to talk casually with people and hear what was on their mind – added to the richness of participant observations.

It was not uncommon for the same clients to return frequently to the clinic, either for methadone/other prescriptions or because they lived in the attached hostel. When the researcher saw an individual she had already spoken with, she made a point of saying hello and asking how that person was getting on. Any new information was recorded after the visit in her field notes. Over the course of her observations there were six individuals who became familiar enough with the researcher to call her by name and sit next to her while they waited to see a doctor.

### Observations

When not in conversation, the researcher observed what was happening around her. Many clients chatted openly amongst themselves or on the phone while waiting. The researcher jotted down relevant concerns that she overheard in her notebook, adding them formally to her field notes later. She also took notes on what she was seeing (e.g., Was anyone sleeping? Limping? How long were clients waiting to be seen? Did people seem in good spirits? Frustrated?).

### Ethical considerations

All participants were 18 years of age or older and have been given a pseudonym. Methods were performed in accordance with the guidelines and regulations outlined in the Declaration of Helsinki and the study passed full ethical review by the University College Dublin Human Research Ethics Committee – [Sciences (HREC-LS)]. Research Ethics Reference Number (REERN): HREC LS-22-42-Ingram-Perrotta. According to recommended guidelines[28, 29], informed oral consent was obtained from all participants who met inclusion criteria and participated in non-recorded, in-depth interviews. When the complex space prevented the obtention of informed consent during participant observations, non-public data were not recorded, and any collected data were analysed and reported only at the general level. To ensure that she did not become emotionally or psychologically distressed when researching sensitive issues, the researcher regularly debriefed with community partners and research colleagues and took breaks between interviews as needed.

### Analysis

Field note data (80 pages, 40,000 words) were coded and analysed using an inductive thematic framework method according to the following recommended stages of trustworthy, thematic analysis [30, 31]:

- **Familiarisation:** Two researchers (CI,IM) familiarized themselves with the data by re-reading the transcripts. Each researcher recorded analytical notes, thoughts, and impressions in the field notes’ margins.
- **Initial coding:** The same researchers independently coded ten pages of field notes line by line, identifying potential themes and subthemes relating to clients’ views on health or healthcare through an ‘open coding’ process. Once results were compared and an initial coding framework constructed, CI and IM coded ten more pages each before further discussing and refining the work. During peer debriefing, the researchers recognized that key themes largely fell under the following categories: self-identified priorities, satisfaction with services, and migrant health. Data from a final theme – barriers and facilitators to addiction recovery – were deemed rich enough to be reported in a separate paper.
- **Applying the thematic framework:** The working thematic framework was systematically applied to all transcripts by CI using Nvivo software V.11. and necessary refinements made until two layers of distinct themes were finalized and approved by all researchers.
- **Charting and interpreting the data:** A matrix was used to summarize data for each code and theme. Connections within and between codes and cases were made in order to fulfil the original research objective of understanding clients’ views on their health and healthcare and highlighting emergent findings generated through inductive analysis.
- **Partner checking:** Community partners were shown a copy of anonymised results and invited to provide feedback on the accuracy of the researchers’ thematic analysis and its implications.

## Results

### Participant Characteristics

Between October 2022 and April 2023, the researcher held in-depth conversations (N=23) and casual conversations (N=15) with 38 clinic attendees and recorded observations for 36 more. Participant characteristics are summarized in Table 1. Conversations and observations were distributed evenly across genders (51% men, 49% women). Most participants were between 20 and 50 years old (N=62, 84%), with only five individuals aged 60 and older. Participants resided in homeless hostels (N=41, 55%) or with family (N=6, 8%). A minority of participants were sleeping rough (N=7, 10%) or were in long term accomodation after experiencing homelessness in their lifetime (N=8, 11%). Of 39 clients with active drug addiction, 32 (82%) were on opioid substitution therapy (OST). Tragically, two participants died of overdose in their hostel during the study observation period. Three clients were waiting to enter a residential treatment programme: 9 clients had completed residential treatment and were in early recovery (<1 year) or sustained recovery (1-5 years) [32] (7 of these continued their OST).

**Table 1.**
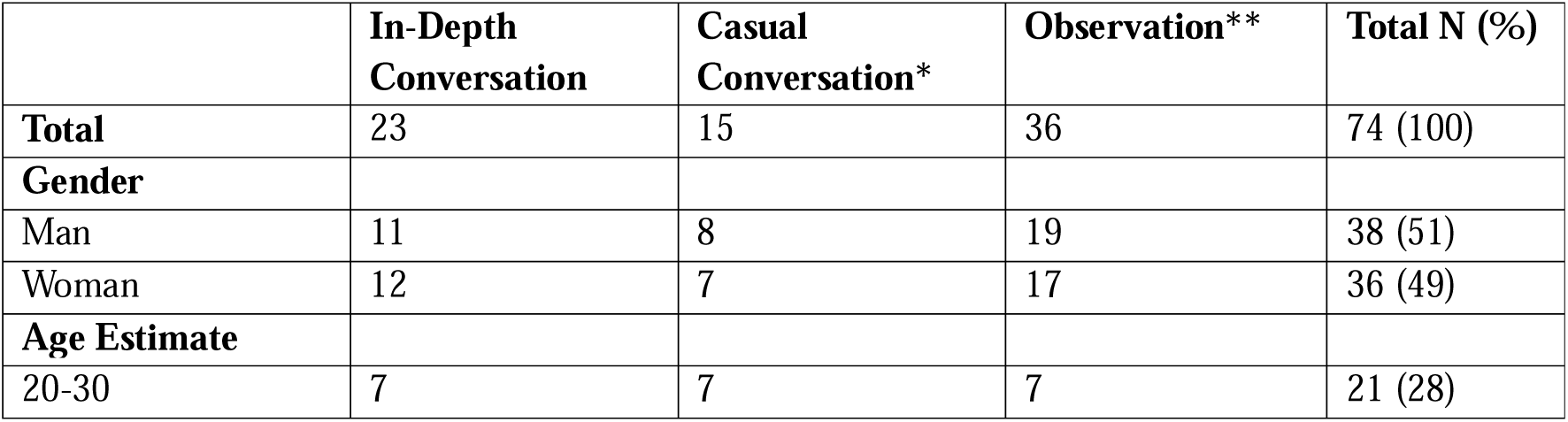
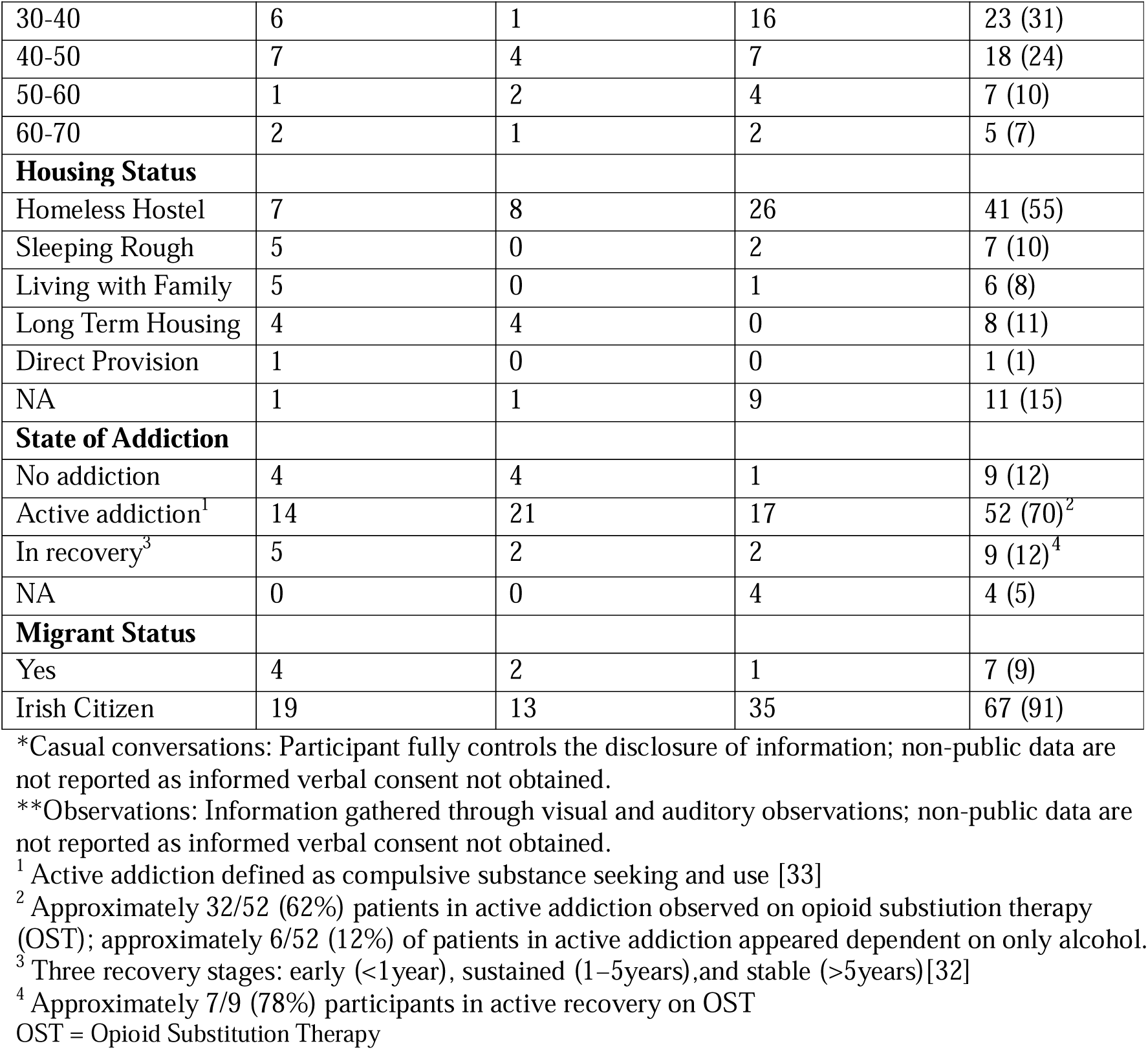
Participant characteristics of 74 clients attending a drop-in primary care and addiction service clinic between October 2022 and April 2023.

In the following sections, we focus on three main themes emerging from our analysis: self-identified priorities, satisfaction with services, and migrant health. The analysis is exemplified with data extracts from the lead researcher’s field notes, making references to participants whose views underpin discursive claims using their pseudonym. Figure 1 provides an overview of the interrelated factors impacting health and wellbeing as identified by study participants.

**Fig 1.**
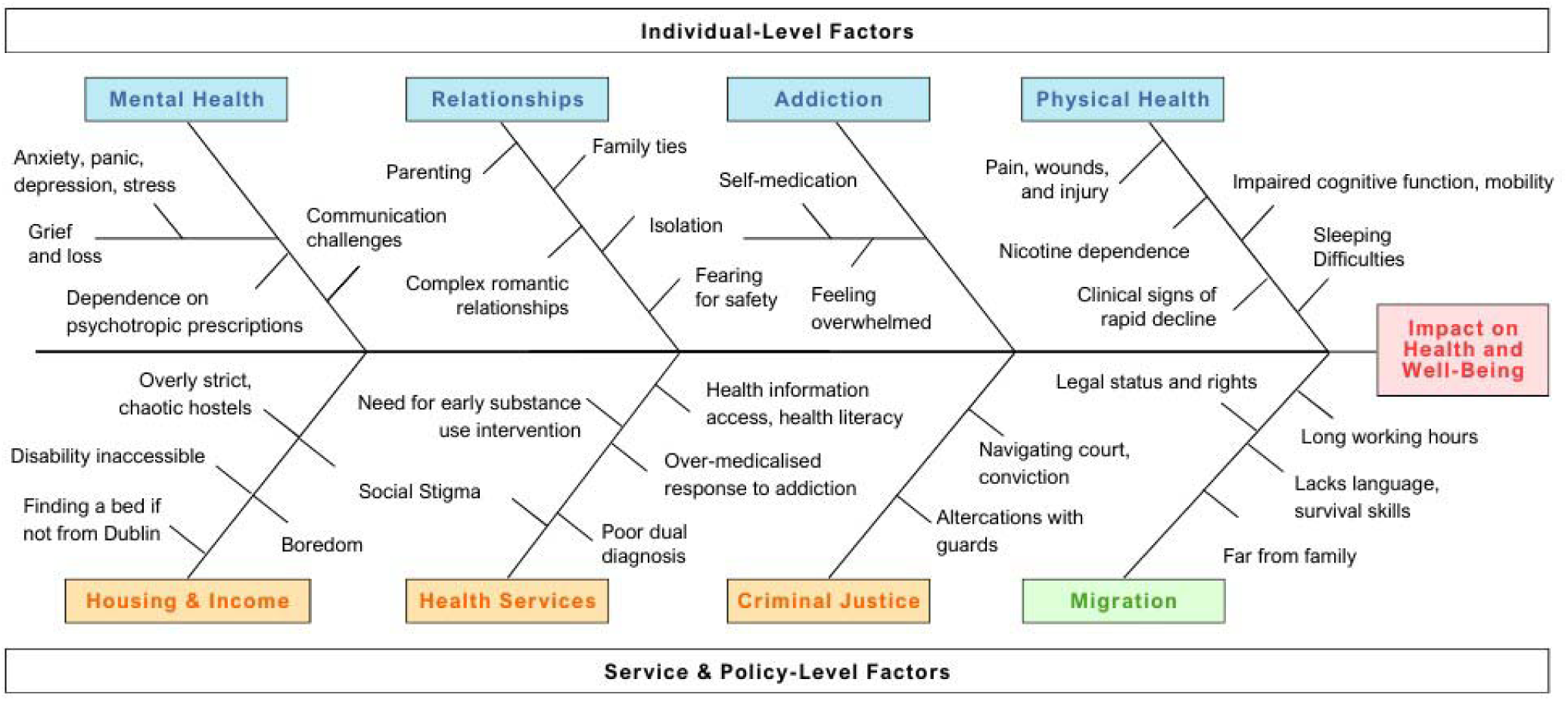
Map of the interrelated factors impacting the health and well-being of people experiencing homelessness attending a Dublin primary care and addiction service: 2022-2023.

### Theme 1 - Self-Identified Priorities

When speaking to the researcher or fellow clinic attendees, clients referred to a number of issues of more pressing concern to them than their long-term physical health or healthcare (Fig 2).

**Fig 2.**
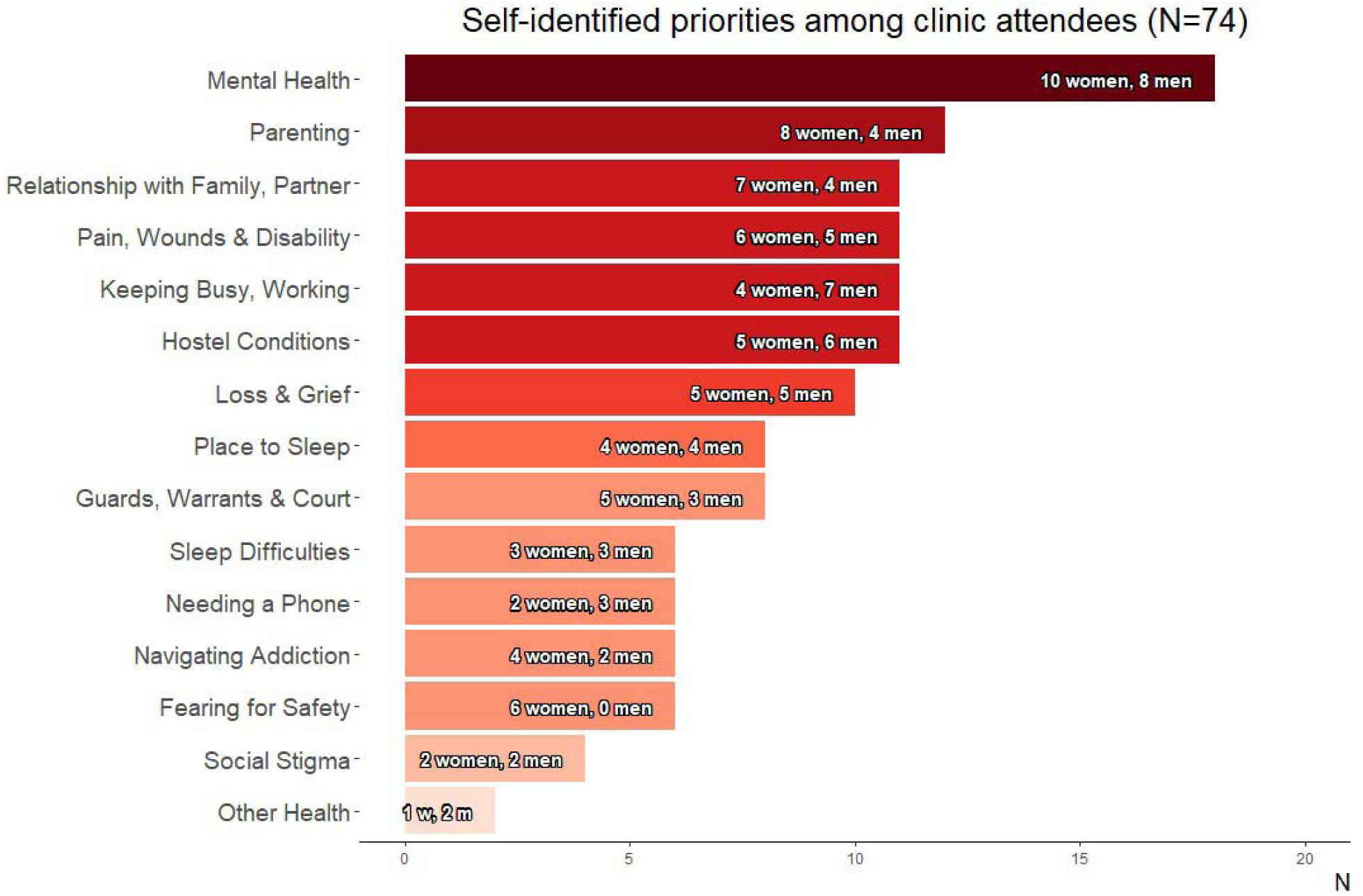
Self-identified priorities among 74 clinic attendees identified through informal interviews and active participant observation: October 2022 to April 2023.

### Mental Health

Clients were most often concerned about the state of their mental health. Those active in their addiction expressed feeling anxious, depressed, panicky, and dependent upon psychotropic medications for those conditions. Often clients needed ‘a smoke’ to calm down in moments of distress or anxiety. Clients mentioned feeling isolated due to their addiction and to the severity of traumatic events experienced. Shame, anger, and stress were expressed frequently. Two individuals mentioned having tried to commit suicide in the past year. One had begun hearing voices though he was not ready to seek help for these.

Mental health conditions presented communication challenges. Some clients remained withdrawn, at times appearing heavily medicated. Others were talkative but difficult to understand due to incoherent ideas. During in-depth conversations, two women expressed frustration at having lost cognitive function due to injuries perpetrated by former partners: *“I can’t focus or communicate things in order anymore” (Eimear)*.

Many clients were grieving the recent loss of a partner, sibling, parent, or close friend. Three participants had lost a family member to suicide in the last year. Another three clients mentioned the distress associated with sharing a room with someone who fatally overdosed in their hostel. One woman explained to a friend, *“I’m the one who found her. I went over to wake her up and saw [she was gone]. All night I slept with her like that” (Observed).* One client was devastated at the guards having taken their dog away while sleeping rough. Others expressed a need for counselling specific to loss, death, and grief:

> *She says her rehab facility charged [nearly 100] euro a week and didn’t provide grief counselling, or any counselling. “I **know** grief”, she says. She is very upset about the loss of her brother. “Broke me heart, broken as it is.” (Teresa)*

### Relationships

Clients spoke often of parenting. Many maintained a positive relationship with their kids and were excited for upcoming outings and time spent together. Others were trying to reduce their substance use so as to reunite with their children. Several women were pregnant or had just had a baby and were enthusiastic about this, though one mother in recovery expressed feeling overwhelmed by a lack of parenting support: *“If there’s one gap in services, it’s social workers for single mothers recovering from addiction whose children are returned to them” (Nora).* Three women mentioned the difficulties of caring for children with special needs (e.g., behavioural issues, autism, attention-deficit/hyperactivity disorder).

Clients were further preoccupied by relations with families and partners. Some were worried about not being able to get in touch with their families and/or from ties being cut. Six clients mentioned their phones being broken, stolen, or confiscated and were upset at having lost numbers and not being able to contact anyone. Others were trying to navigate visiting and caring for elderly or sick relatives. Instances of methadone prescriptions being revised to accommodate these visits were observed by the researcher; something that clients were grateful for each time. Several clients were struggling with recent breakups of long-term relationships. Women noted trying to navigate complex intimate relationships including those with violent partners; with partners whose behaviour had recently changed in relation to their drug use; with partners who remained in addiction as they tried to recover; and with partners set to be released from prison.

### Safety

Tying in with relationships was the issue of personal safety. Women told the researcher about current threats against their lives by abusive ex-partners:

> As we’re speaking, she starts to break down. She tells me that the man who violently assaulted her just got out of prison on compassionate grounds. “Where is the compassion for me?!” she says. She saw him on the street that morning and had to get off the bus to get sick. (Eimear)

Clients commented on how difficult it is to be a woman in homelessness: *“If you have drugs in your bra or panties, those guys aren’t afraid to come and find it there” (Cara).* One woman had been asked to leave a domestic violence shelter when her abuser found out where she was staying. The researcher asked her how she stayed safe after leaving, at which point the woman took out her removable partial denture to reveal a missing tooth. Another client explained that despite being able to stick up for anyone else, she couldn’t seem to find the same anger or strength to stick up for herself in abusive situations.

### Housing and Income

Some clients were preoccupied with finding a bed for the night. On three separate occasions clients spent long stretches on hold with the ‘Freephone’, the number to call to access emergency accommodation in Dublin. Frustrations ensued when the phone line cut off after an hour and the process had to be repeated. Another older client could not access a bed for the night because he was not from the city. He had been sleeping rough for months and was struggling to carry his belongings around while using crutches for a hip injury. Several clients had been waiting years to access more permanent social housing.

Clients able to access beds in hostels were frustrated by the prevalence of drug use within them. One woman waiting for a spot in residential treatment relied on smoking weed to *“at least be doing something” (Ailis)* while her roommates used. For another individual detoxing from alcohol, the lack of private, supportive environment was proving difficult as he went through severe withdrawals. Strict rules (e.g., no paracetamol allowed in hostel) and condescending, inexperienced staff were of concern. Upon entering homelessness, one man with severe mobility impairments had been assigned a hostel bed on the third floor of a building with no elevator.

Boredom brought on by limited resources and mobility was frustrating to those who found themselves idling all day in homeless accommodation. Some clients were unable to work due to disability or immigration status; others did not want to seek employment for fear of losing disability payments. For clients who were working, satisfactory job choices could be limited due to scheduling conflicts with methadone prescriptions or failing to obtain garda vetting.

Social stigma impeded clients’ ability to manage their day-to-day affairs. When interacting with social welfare or other services, several told the researcher about experiences of being hung up on or ignored.

> Sean tells me that he tried to ring social services this week and was hung up on. He shows me a card with a list of numbers to call – at least 10 – it’s very confusing. He says when he finally got someone on the phone he said, six times, “Please don’t hang up on me”, but the person did. He tried to set up an account this week at the Credit Union. The girl behind the counter kept asking him questions that didn’t make sense for someone in homelessness. He says there’s a lot of people in those type of positions that don’t know what to make of him. (Sean)

### Physical Health

Despite being in a general practice, only three clients mentioned physical health conditions beyond pain, sleep, or disability. One man proudly informed the researcher that he’d never missed taking his HIV tablets: *“don’t forget that undetectable means untransmissible!” (Tyler).* Another client was trying to get their teeth fixed and a third mentioned dealing with bladder incontinence. Clients often complained of sleep deprivation due to behaviours of roommates in hostels, conditions while sleeping rough, and mental health conditions (e.g., panic, anxiety, post-traumatic stress disorder). The researcher frequently observed clients sleeping in the waiting room.

Not infrequently, clients reported to the GP with wounds self-treated with toilet paper, cellotape, plastic shopping bags, or superglue. One man who came in for *“just a knife wound …nothing too painful” (Gavin),* said he would not present to hospital for his injury to avoid having to report the perpetrator and becoming a target when back on the street. Pain was a common theme. Many clients came into the clinic limping or wincing as they sat and stood. Others in varying stages of alcohol detoxifcation experienced tremors of the hands and face as they waited. More vocal clients would call out the extent of their pain if frustrated by wait times. Over the seven month observation period, the researcher observed the rapid decline in health of several individuals who grew more frail, thin, and withdrawn over time before – in one instance – passing away.

### Guards, Warrants, and Court

Clients mentioned negative interactions with the guards involving having belongings taken, being called a junkie, and *“getting battered.”* Clients were preoccupied with upcoming court dates (as many as 12) and how going to prison would interfere with entering residential treatment or taking care of their children. One man noted that he had to be careful out walking around due to the warrants out for his arrest *“for small things, stealing the odd can” (Ryan).* The researcher noticed a tendency for clients to take accountability for their actions: *“It’s fair really. I’ve caused a lot of trouble from the drinking” (Leonard)*.

### Navigating Addiction

Six clients active in addiction expressed feeling overwhelmed by their substance use. Four of these individuals vocalized wanting help, while two others did not seem ready to accept it: *“there’s just no way I’m getting through the next few days sober” (Pamela).* One man had been working on reducing his methadone dose for months to be able to enter treatment on a particular Friday. When told that he would need to wait until the Monday he was crushed; he told a member of staff, *“if I’m going to use, today’s going to be the day” (Observed)*.

### Theme 2 - Satisfaction with Services

#### Drop-In Primary Care & Addiction Service

The clinic itself was a social environment, with noticable camraderie between staff and clients and amongst groups of acquaintances. Many clients came with a friend or made plans to meet up later. Smoking was a big part of the clinic social scene. Clients frequently shared tobacco and congregated outside to smoke and chat. Word travelled in the waiting room about the quality of various doctors, nurses, addiction services, hospitals, and hostels across the city. The researcher heard one woman dissuade a friend from entering a particular stabilisation facility:*“that place is a kip” (Observed).* Tips and tricks of the trade were frequently shared such as how to access a bed for the night or how to ask for sleeping pills.

In terms of complaints, the clinic grew more crowded over the researcher’s observation period. Some clients were frustrated by wait times and frequently took these frustrations out on staff. Others were frustrated by drug dealing and use in close proximity or by occasional chaotic situations in the waiting room. However the majority of clients, when asked about their healthcare, reported feeling satisfied with the services they were receiving in the clinic. This mostly came down to the care provided by the medical professionals and operations staff. One client explained that it was the first time she felt that she could be honest with her doctor about her drug use: *“It feels really good to be trusted. It feels good that people know my name” (Róisín)*.

#### Information

Clients noted instances where they lacked desired information regarding their healthcare. One man – unaware that Librium was not meant to be taken with alcohol – said that he had taken a packet of tablets, drunk a bottle of liquor, and wound up in an altercation with the guards. One man didn’t understand why he’d been asked to return to get his bloods done multiple times. Another man completing a benzodiazepine detox wasn’t sure why he was still being handed stacks of pills each visit. He complained to a friend, *“I’m trying to get off this stuff, I don’t want to be on these forever.” (Observed)*

Clients also mentioned a desire for proactive outreach for those in addiction. One man wished that more of an effort was made to go into communities and target individuals in early stages of substance use: *“Make sure to tell the doctor that. You have to get to the young people. It’s really important.” (Timmy)* Another hoped to see more information and outreach regarding non-medicalized options for recovery:

> Lorcan says the pain of facing your trauma is not as bad as the pain of addiction. He wishes people knew this. When GPs hear somebody say ‘I’m fed up. I can’t do this anymore’, he wishes they would point them towards stabilisation, residential treatment, 12-step. He says that a lot of people, even GPs, don’t know about the services available. They don’t know that there is a way to face the pain besides drugs and alcohol. In addiction, those seem like the only option. He wants people to know that it’s not. (Lorcan)

#### Opioid Substitution Therapy

Several clients noted frustrations with methadone prescribing:

> Jack says that if he could change one thing, it would be how hard it is to get off methadone. He explains that now, a person starts on a lower dose until, say a few weeks in, they use street drugs. The doctors then raise the methadone dose. To him, this doesn’t make sense. If a person was having withdrawals so severe that they needed to take illicit drugs in addition to their methadone, that would have happened early on, not several weeks in. He doesn’t think raising the dose is the solution because it’s very hard to get off. (Jack)

Two clients said that when they expressed a need for help with their addiction, their private GP had only wanted to talk about methadone without mentioning other options. One found a new GP who was willing to follow the client’s own plan for recovery. Conversely, two women who had been on methadone for years mentioned that it was going well. Across her visits, the researcher observed clear improvements in stability in those on lower doses of methadone compared to those just starting.

The topic of convenience came up. For one client, picking up methadone prescriptions clashed with working hours: *“I’ve a key worker that can pick it up for me but she’s not always around” (Timmy).* Another individual missed her methadone appointment, ran out over the weekend, and had resorted to taking street drugs. Two women on suboxone liked its convenience - *“I can carry it around in my purse” (Aileen)* - but one said she could have used extra support when transitioning from methadone to suboxone due to the severity of withdrawals. On one clinic visit, the researcher overheard two clients discussing how they were being prescribed methadone for pain but didn’t want it: *“I’ve never taken a drug in me life, I don’t want that stuff” (Observed)*.

#### Mental Healthcare

Seven clients talked about their experiences with mental healthcare. One man explained his issue with dual diagnosis:

*Jack says that he went to a psychiatrist a day that he was suicidal and was sent back to the streets with two pills, one for the night and one for the morning. He took both immediately. The psychiatrist said that the suicidal thoughts were linked to his drug use and not his mental health. ‘That doesn’t make sense because I’ve been using drugs since I was a teenager but I’m only suicidal since my partner kicked me out.’ (Jack)*

Another woman faced a similar dilemma, *“I can’t get psych medication until my addiction is sorted but the more I get my addiction sorted, the more it’s bringing up old emotional stuff that I need help dealing with” (Alannah).* In other instances, mental health was responded to as a law enforcement issue as demonstrated by this exchange between a client and the researcher:

*She tells me of one episode of psychosis so bad that she ended up covered in blood running in and out of cars. The police came but didn’t call the ambulance. ‘Why?’ I ask. She laughs, ‘You think they’d come for me?! Having a psychosis, supposedly on drugs? That could take ages.’ Instead, she was put into a jail cell until she was feeling well enough to leave. The police know her. ‘They did charge me,’ she says, ‘obstruction of traffic.’ (Nora)*

One woman had been seeing a therapist based in her hostel but had recently changed housing. Another said that her issues and experiences had been too severe for the domestic violence counsellor she had seen in the past: *“I needed more help than she could give me” (Eimear).* There were positive stories. A woman who had been assualted got a letter from the GP to see a psychiatrist and felt better being able to talk to someone. Other clients, when offered mental health support via the clinic’s drop-in psychotherapist, passed on this:*“Oh you know me, once you got me talking I just wouldn’t stop” (Alannah)*.

### Theme 3 - Migrant Health

Migrant participants dealt with a different set of competing priorities unrelated to substance misuse. The researcher spoke with four African individuals attending the clinic who had obtained European citizenship in other countries but – due to precarious tenancy agreements and lack of affordable housing – were living in temporary homeless accommodation in Ireland. For these individuals, primary concerns related to working long hours, navigating long commutes to work, and applying for the Housing Assistance Payment (HAP). Other, asylum seekers faced unique challenges relating to traumatic, months-long journeys; lack of beds in Direct Provision and having to sleep rough upon arrival in Ireland; not being allowed to work before the processing of the asylum application and having to survive on a weekly allowance of €38; and fearing for the safety of family back home. These challenges impacted health. One man noted that despite being pre-diabetic, he’d resorted to eating lots of candy since arriving in Ireland as a small comfort.

All migrant participants were trying to navigate living far from family and legally bringing their spouses and children to Ireland. Language barriers were present. On multiple occasions, groups of migrants attended the clinic with a ‘spokesperson’ (usually a friend) to serve as an interpreter. While many of the Irish clients seemed familiar with one another, groups of clients from other countries typically remained to themselves and spoke in their respective languages. Unprompted, a man experiencing homelessness and addiction who had grown up in Ireland described how refugees may lack the skills necessary for surviving on the streets:

> *Gavin said the refugee recognized him on the street and asked for help finding some cardboard and a marker to write up a sign saying that he [the refugee] was from Ukraine and homeless. Gavin found the supplies and started helping him ask people for money. He says the man was shocked when they made about* €*10 in five minutes. He explained that refugees coming in don’t have the street skills he’s had to use all his life (Gavin)*.

## Discussion

In light of a lack of evidence in Ireland on how PEH would like their own healthcare, this study used an ethnographic approach to explore the perspectives of people attending a walk-in primary care and addiction service on their health and healthcare needs. Clients were overwhelmingly concerned about the state of their mental health, expressing pervasive panic, anxiety, depression, and complex mental health disorders that negatively interacted with all other aspects of their lives. The issues most harmfully impacting mental health included isolation from family, loss of loved ones, proximity to death, and fears for safety after experiences of domestic, sexual, and gender-based violence. The factor most positively impacting mental health was meaningful, positive relationships, particularly with one’s children and family, though supportive friendships and relationships with healthcare providers also played a constructive role. Despite being in pain, many clients were not ready to consider addressing the underlying causes of their mental distress while still navigating the complexities of precarious housing, relying heavily instead on self-medication with street drugs and prescribed psychotropics.

Mental health concerns are internationally recognised as being of high priority to homeless service users [34–41]. Depression [34–39], anxiety [34–36], post-traumatic stress [35], and general emotional distress [40] are widespread in homelessness [36]. Our finding that clients with bi-morbid mental illness and addiction struggle to access mental healthcare is not unique to Ireland. Evidence from the UK, Australia, and Canada outlines similar issues with dual diagnosis whereby individuals are expected to be substance-free before accessing mental health supports despite relying on those very substances to block out traumatic experiences and memories [35, 37, 42]. Homeless service users interviewed in similar studies have expressed a need for more available and accessible mental health and psychiatry services [35, 41, 43]. Despite evidence from Ireland that there is a critical shortage of mental healthcare beds and professionals [11], clients we spoke with more often expressed concerns about the acceptability rather than the availability of existing supports. In some instances, this was related to the internalised belief of being beyond help. Other times, in line with previous findings, service users desired targeted help to work through loss and grief [34, 35]. One recommended response would be to create space and opportunity for memorial services within housing services or public settings to reduce social exclusion and disenfranchised grief, and provide a form of advocacy [44]. In Seattle, Homeless Housing First residents recommend integrating grief counselling into single-site or shelter settings where residents who live in close proximity and share communal spaces may be deeply affected by the death of a fellow resident [45].

Service users frequently mentioned the role that close relationships played in relation to their mental health and wellbeing. Researchers have outlined how the integration of homeless individuals into social networks contributes to better physical and mental health status and lower likelihood of victimisation [46]. We observed how the community health clinic fostered social support by providing a space for people to come together to converse. This phenomenon of the clinic waiting room as a social hub for those in homelessness was recently identified in Canada [47]; service users frequently attend walk-in services with their peers, look out for each other, and encourage others to seek help to feel better. We witnessed firsthand how supportive relationships with service providers and staff that include humour and compassion helped to rebuild service users’ self-worth and confidence [43]. Moving forward, our findings reinforce the need for more and/or better-publicised social activities that provide meaningful ways to combat boredom and strengthen bonds formed while in homelessness. Successful examples from the literature include opportunities to come together for work, education, volunteering, and recreation [43]. Despite many female service users being in intimate relationships, we did not hear of (nor did we find evidence of) women seeking and finding social support through their partners. Instead, relationships with family and children stood out as essential and evoked strong emotions [34, 48, 49]. The need for more childcare services and concerns about children’s behaviours, mental health, and education while in homelessness are longstanding [49]. Our finding that social care tapers away prematurely once parents enter recovery and find more stable housing has not been widely reported and merits further exploration.

Another layer of complexity was added for women we spoke with who – in addition to navigating mental health concerns, fear, and family responsibilities – were dealing with loss of cognitive function due to traumatic injury. Unhoused women internationally have identified the impact of injuries and trauma stemming from domestic violence as their priority health concern [34]. That women in homelessness present to community health clinics after experiences of sexual, domestic, or gender-based violence creates opportunity in crisis to link in with refuge spaces and psychological supports provided they are available. Despite a chronic shortage of both in Ireland [11, 50], we noted that clinic staff were generally apprised of clients’ safety concerns and signposted existing supports when possible. Recommendations for improving responses and interventions for homeless women with complex needs in future include acknowledging and addressing structural inequalities, building trust, attending to safety, and improving immediacy and pathways [51]. Beyond these, we recommend exploring the feasibility and acceptability of adapting rehabilitation interventions for traumatic brain injury (TBI) for women in homelessness based on frustrations expressed regarding memory loss and inability to focus. Promisingly, programmes and interventions for TBI specific to those in homelessness exist and have shown promise in the US and Canada [52].

Studies from the US, New Zealand, Toronto and Ireland have identified untreated substance use disorder as an unmet health need amongst PEH [36, 38, 40, 41, 53], calling for greater awareness surrounding the range of supports available [53]. This matches the desire expressed by several of our participants for GPs to promote more non-medicalised treatment options. However, during peer debriefing, our clinic partners explained that GPs may hesitate to promote residential addiction treatment services that provide only a temporary fix and create risk of relapse and overdose upon discharge into homelessness [54, 55]. It may also be unfair to promote non-medicalised supports that are in short supply. Current wait times for someone in homelessness to enter residential treatment in Ireland range from 3 to 6 months; a period during which many will be staying in shared accommodation surrounded by individuals using drugs while trying to reduce their own substance use to meet strict entry requirements for residential treatment [11]. The challenging nature of these circumstances raises the question of how health practitioners can balance parallel responsibilities to maintain non-maleficence from prematurely promoting recovery while also respecting patient autonomy to learn about and choose treatment options in line with their goals. One compromise may be to mention avenues for clients to participate in meaningful social activities and available community-based treatments [56] with a view that regardless of substance use outcomes, social support and perceived choice are good for health [46, 57]. Explaining in part the strong and differing opinions we heard on methadone, research shows that service users with opioid use disorder generally have limited understanding of the chronicity of their illness, the likelihood for long-term OST or the rationale for using methadone as a medical intervention in their disease management [58]. As such, GPs might have regular conversations with patients on OST to create realistic expectations, discuss the rationale behind the treatment, and ascertain if any changes in medication are desired that might be safely made.

Unrelated to substance use, increasing rates of migrants and asylum seekers are entering Ireland straight into homelessness [11]. While many of the concerns we heard were common across homeless cohorts (e.g., dealing with grief and trauma, separation from children and family, finding a place to sleep), language barriers and limited rights to welfare, housing, and medical care were specific to clients recently arrived in Ireland. Clinic staff attempted to direct these individuals to targeted non-governmental organisations, however they recognised – as we heard firsthand – that supports for those entering the country into homelessness are extremely limited. Where supports do exist, data show that refugees and other migrants often lack information about them, finding them difficult to access and navigate [36, 59]. We did not get a sense that migrant clients benefited from the social atmosphere of the community health clinic in the same way as their Irish counterparts. Instead, its ease of access is likely why those who had obtained EU citizenship and legal employment continued to attend the service despite being eligible for / encouraged to use mainstream primary care. It was interesting to note that despite high rates of traumatic experiences, migrant clients less frequently expressed concern over their mental health, begging the question of how psychosocial supports for those in homelessness might be viewed / prioritised differently across different cultures. As a whole, these preliminary findings underline the importance of assessing the unique health needs specific to individuals experiencing homelessness after migrating from diverse countries and conflicts.

Finally, in terms of physical health, our finding that preventive healthcare was deprioritized in our study cohort in favour of managing competing priorities has been widely documented [4, 35, 37], as have self-identified concerns about sleep [34, 41], chronic pain [34, 39, 60], and loss of both physical and cognitive function due to injuries resulting in/from homelessness [34, 39, 41]. Encouragingly, service users did not mention cost as a barrier to care as has been noted in other countries [34, 47], nor did the issue of food insecurity come up [37, 40]. Homelessness is associated with high rates of tobacco smoking [35] and we witnessed a strong reliance on tobacco as a small comfort in the face of stress and sadness. Primary prevention is of course a vital component towards achieving health equity. However, the anecdotes we heard regarding the accessibility and quality of temporary housing drive home the impracticality of expecting individuals to focus on improving any aspect of their health before they have safe and stable accommodation.

### Limits

This study was located in one clinic providing accessible, walk-in primary care and addiction services which impact our findings in several ways. First, findings capture the views of a population linked with health services. While we did identify crossover with views expressed by homeless service users in Canada, the US, the UK, Sweden, Norway, Australia, New Zealand, and South Africa [34, 35, 37–43, 47–49, 53, 60, 61], our findings may not be transferrable to individuals who are not accessing care nor to locations where access to primary care is more limited for those in homelessness. Second, in some instances clients we spoke with may have been hesitant to share views on the care they were receiving in the same clinic where interviews were taking place. While not recording the interviews allowed us to capture organic and informal conversations, we may have missed certain nuances of clients’ perspectives in the writing up of field notes. As well, some clients were hard to understand due to mumbling, slurring, or incoherent ideas and not all of what they shared could be documented. This served as a poignant reminder to the researchers that often, individuals who need support the most are less likely to be able to communicate their needs and/or advocate for themselves.

## Conclusions

Using an ethnographic approach, this study sought to explore the views of people attending a walk-in primary care and addiction service on their own health and healthcare needs. Many of the priority concerns identified by homeless clients are not dissimilar from those of the general population. Clients are concerned about their mental health, maintaining ties with their children and families, navigating complex romantic relationships, finding meaningful activities, and feeling better physically. The difference is the frequency and severity of these concerns observed both prior to, and during experiences of homelessness exacerbated by disproportionate exposure to loss, fear, injury, pain, disability, fatigue, and isolation.

An encouraging finding is that clients taking part in this research are satisfied with their ability to access primary care and harm reduction services in a social environment where positive exchanges with friends and providers take place. Conversely, barriers to accessing mental health and addiction services persist, including the internalised belief that one is beyond help, lack of access to information on available services and their entry requirements, and lingering stigma within a health system that treats addiction as separate to health. Recommendations for practice include holding more regular and open conversations with homeless clients during primary care consultations about the care they are receiving, its rationale, and whether or not changes are desired that can be safely made. In consultation with service users and providers, policymakers should prioritise expanding access to refuge spaces, parenting supports, and residential stabilisation, treatment, step-down, and aftercare beds for individuals experiencing homelessness. The health needs of migrants and asylum seekers entering homelessness in Ireland are urgent and, as they are not yet widely understood, should be prioritised in future research.

## Data Availability

All relevant data are within the manuscript and its Supporting Information files.

## Acknowledgements

We would sincerely like to thank community health clinic partners for contributing their practical and clinical expertise to this project and facilitating the project space and participant recruitment. We acknowledge that this project was made possible in large part thanks to the trust built over time between clinic staff and clients which allowed us to be accepted more quickly into the space as research colleagues.

## Notes

### Competing Interest Statement

The authors have declared no competing interest.

### Funding Statement

The author(s) received no specific funding for this work.

### Author Declarations

The Human Research Ethics Committee - Sciences of University College Dublin gave ethical approval for this work (Research Ethics Reference Number: HREC LS-22-42-Ingram-Perrotta).

## References

[1] Fazel S, Geddes JR, Kushel M. The health of homeless people in high-income countries: descriptive epidemiology, health consequences, and clinical and policy recommendations. The Lancet 2014; 384: 1529–1540.

[2] Ingram C, Buggy C, Elabbasy D, et al. Homelessness and health-related outcomes in the Republic of Ireland: a systematic review, meta-analysis and evidence map. J Public Health (Berl). Epub ahead of print 1 June 2023. DOI: 10.1007/s10389-023-01934-0.

[3] World Health Organization, Pan American Health Organization. Health Equity, https://www.paho.org/en/topics/health-equity (2023, accessed 20 September 2023).

[4] O’Carroll A, Wainwright D. Making sense of street chaos: an ethnographic exploration of homeless people’s health service utilization. Int J Equity Health 2019; 18: 113.

[5] Allen M, Benjaminsen L, O’Sullivan E, et al. Trends in homelessness in Denmark, Finland and Ireland. In: *Ending Homelessness?* Bristol University Press, 2020, pp. 73–102.

[6] Dublin Regional Homeless Executive. *Monthly Report to Dublin City Councillors on Homelessness*, chrome-extension://efaidnbmnnnibpcajpcglclefindmkaj/https://www.homelessdublin.ie/content/files/Homeless-update-39-May-2023.pdf (December 2023).

[7] Dublin Regional Homeless Executive. Week long assessment of rough sleepers in the Dublin Region, chrome-extension://efaidnbmnnnibpcajpcglclefindmkaj/https://www.homelessdublin.ie/content/files/Spring-2023-Count-of-Rough-Sleepers.pdf (December 2023).

[8] REDC, Simon Community. Simon Community - Hidden Homelessness Poll, https://www.simon.ie/wp-content/uploads/2022/09/Simon-Communities-of-Ireland_Hidden-Homeless-Poll.pdf (2022, accessed 5 January 2023).

[9] Ena Lynn, Joan Devin, Sarah Craig, et al. Deaths among people who were homeless at the time of death in Ireland, 2019. National Drug-Related Deaths Index, Health Research Board, chrome-extension://efaidnbmnnnibpcajpcglclefindmkaj/https://www.hrb.ie/fileadmin/2._Plugin_related_files/Publications/2023_Publications/12726_HRB_Deaths_among_homeless_people_Statlink_11_FA_WEB.pdf.

[10] O’Sullivan K. *Primary care access for homeless people: identifying best practice using Merchants Quay Ireland as a model of primary care provision*. Working Paper, Combat Poverty Agency (CPA), https://www.lenus.ie/handle/10147/106646 (September 2008, accessed 8 September 2023).

[11] Ingram C, MacNamara I, Buggy C, et al. Priority healthcare needs amongst people experiencing homelessness in Dublin, Ireland: A qualitative evaluation of community expert experiences and opinions. *PLOS ONE* 2023; 18: e0290599.

[12] Ivers J-H, Zgaga L, O’Donoghue-Hynes B, et al. Five-year standardised mortality ratios in a cohort of homeless people in Dublin. BMJ Open 2019; 9: e023010.

[13] Ingram C, Roe M, Downey V, et al. Exploring key informants’ perceptions of Covid-19 vaccine hesitancy in a disadvantaged urban community in Ireland: Emergence of a ‘4Cs’ model. Vaccine 2023; 41: 519–531.

[14] O’Carroll A, Duffin T, Collins J. Harm reduction in the time of COVID-19: Case study of homelessness and drug use in Dublin, Ireland. International Journal of Drug Policy 2021; 87: 102966.

[15] Healthy London Partnership. Homeless Health Needs Assessment toolkit: Assessing single homeless people’s health needs in your locality. 2018.

[16] Cain CL, Orionzi D, O’Brien M, et al. The Power of Community Voices for Enhancing Community Health Needs Assessments. Health Promotion Practice 2017; 18: 437–443.

[17] Delivering the Goods, Showing Our Stuff: The Case for a Constructivist Paradigm for Health Promotion Research and Practice - Ron Labonte, Ann Robertson, 1996, https://journals.sagepub.com/doi/epdf/10.1177/109019819602300404 (accessed 4 October 2023).

[18] Cubellis L, Schmid C, von Peter S. Ethnography in Health Services Research: Oscillation Between Theory and Practice. Qual Health Res 2021; 31: 2029–2040.

[19] Berry NS, McQuiston C, Parrado EA, et al. CBPR and Ethnography: The Perfect Union. In: Methods for Community-Based Participatory Research for Health. San Francisco, California: Jossey-Bass, 2013.

[20] Eng E, Strazza K, Rhodes S, et al. Insiders and outsiders assess who is ‘The Community’: Participant observation, key informant interview, focus group interview, and community forum. In: Methods for Community-Based Participatory Research for Health. San Francisco, California: Jossey-Bass, 2013.

[21] Sarah J. Tracy. Qualitative Research Methods: Collecting Evidence, Crafting Analysis, Communicating Impact. 2nd ed. Wiley Blackwell, https://www.wiley.com/en-us/Qualitative+Research+Methods%3A+Collecting+Evidence%2C+Crafting+Analysis%2C+Communicating+Impact%2C+2nd+Edition-p-9781119390800 (2020, accessed 22 March 2023).

[22] Goedhart NS, Pittens CACM, Tončinić S, et al. Engaging citizens living in vulnerable circumstances in research: a narrative review using a systematic search. Research Involvement and Engagement 2021; 7: 59.

[23] Swain JM, Spire ZD. The Role of Informal Conversations in Generating Data, and the Ethical and Methodological Issues. Forum Qualitative Sozialforschung / Forum: Qualitative Social Research; 21. Epub ahead of print 28 January 2020. DOI: 10.17169/fqs-21.1.3344.

[24] Government of Ireland. Housing Act. *electronic Irish Statue Book (eISB)*, https://www.irishstatutebook.ie/eli/1988/act/28/enacted/en/print#sec2 (1988, accessed 23 November 2022).

[25] Michael Quinn Patton. *Qualitative Research & Evaluation Methods*. 4th ed. Sage, https://us.sagepub.com/en-us/nam/qualitative-research-evaluation-methods/book232962 (2002, accessed 23 March 2023).

[26] McLean K. Needle exchange and the geography of survival in the South Bronx. Int J Drug Policy 2012; 23: 295–302.

[27] Bungay V, Johnson JL, Varcoe C, et al. Women’s health and use of crack cocaine in context: Structural and ‘everyday’ violence. International Journal of Drug Policy 2010; 21: 321–329.

[28] Zahle J. Privacy, Informed Consent, and Participant Observation. Perspectives on Science 2017; 25: 465–487.

[29] Guest G, Namey EE, Mitchell ML. *Collecting Qualitative Data: A Field Manual for Applied Research*. SAGE Publications, Ltd. Epub ahead of print 2013. DOI: 10.4135/9781506374680.

[30] Nowell LS, Norris JM, White DE, et al. Thematic Analysis: Striving to Meet the Trustworthiness Criteria. Int J Qual Methods 2017; 16: 1–13.

[31] Gale NK, Heath G, Cameron E, et al. Using the framework method for the analysis of qualitative data in multi-disciplinary health research. BMC Med Res Methodol 2013; 13: 117.

[32] Martinelli TF, Nagelhout GE, Bellaert L, et al. Comparing three stages of addiction recovery: long-term recovery and its relation to housing problems, crime, occupation situation, and substance use. *Drugs: Education*, Prevention and Policy 2020; 27: 387–396.

[33] Abuse NI on D. Drug Misuse and Addiction. *National Institute on Drug Abuse*, https://nida.nih.gov/publications/drugs-brains-behavior-science-addiction/drug-misuse-addiction (--, accessed 15 September 2023).

[34] Sutherland G, Bulsara C, Robinson S, et al. Older women’s perceptions of the impact of homelessness on their health needs and their ability to access healthcare. Australian and New Zealand Journal of Public Health 2022; 46: 62–68.

[35] Mc Conalogue D, Maunder N, Areington A, et al. Homeless people and health: a qualitative enquiry into their practices and perceptions. Journal of Public Health 2021; 43: 287–294.

[36] O’Donnell P, Tierney E, O’Carroll A, et al. Exploring levers and barriers to accessing primary care for marginalised groups and identifying their priorities for primary care provision: a participatory learning and action research study. International Journal for Equity in Health 2016; 15: 197.

[37] Campbell DJT, O’Neill BG, Gibson K, et al. Primary healthcare needs and barriers to care among Calgary’s homeless populations. BMC Family Practice 2015; 16: 139.

[38] Bukowski K, Buetow S. Making the invisible visible: A Photovoice exploration of homeless women’s health and lives in central Auckland. Social Science & Medicine 2011; 72: 739–746.

[39] Seager JR, Tamasane T. Health and well-being of the homeless in South African cities and towns. Development Southern Africa 2010; 27: 63–83.

[40] Daiski I. Perspectives of homeless people on their health and health needs priorities. Journal of Advanced Nursing 2007; 58: 273–281.

[41] Thorndike AL, Yetman HE, Thorndike AN, et al. Unmet health needs and barriers to health care among people experiencing homelessness in San Francisco’s Mission District: a qualitative study. BMC Public Health 2022; 22: 1071.

[42] Bennett-Daly G, Maxwell H, Bridgman H. The Health Needs of Regionally Based Individuals Who Experience Homelessness: Perspectives of Service Providers. International Journal of Environmental Research and Public Health 2022; 19: 8368.

[43] Diduck B, Rawleigh M, Pilapil A, et al. Mental health needs of homeless and recently housed individuals in Canada: A meta-ethnography. Health & Social Care in the Community 2022; 30: e3579–e3592.

[44] Monk J, Black J, Carter RZ, et al. Bereavement in the context of homelessness: A rapid review. Death Studies 2023; 0: 1–10.

[45] Mackelprang JL, Clifasefi SL, Grazioli VS, et al. Content Analysis of Health Concerns among Housing First Residents with a History of Alcohol Use Disorder. Journal of Health Care for the Poor and Underserved 2021; 32: 463–486.

[46] Hwang SW, Kirst MJ, Chiu S, et al. Multidimensional Social Support and the Health of Homeless Individuals. J Urban Health 2009; 86: 791–803.

[47] Paradis-Gagné E, Jacques M-C, Pariseau-Legault P, et al. The perspectives of homeless people using the services of a mobile health clinic in relation to their health needs: a qualitative study on community-based outreach nursing. J Res Nurs 2023; 28: 154–167.

[48] Kneck Å, Klarare A, Mattsson E, et al. Reflections on health among women in homelessness: A qualitative study. Journal of Psychiatric and Mental Health Nursing 2022; 29: 709–720.

[49] Gelberg L, Browner CH, Lejano E, et al. Access to Women’s Health Care: A Qualitative Study of Barriers Perceived by Homeless Women. Women & Health 2004; 40: 87–100.

[50] Bourke AM. Domestic Violence and Women in Homelessness. *Simon Communities in Ireland*, https://www.simon.ie/domestic-violence-and-women-in-homelessness/ (2022, accessed 18 December 2023).

[51] Morton S, MacDonald S, Christophers L. Responding to Women with Complex Needs Who Use Substances: A briefing paper. University College Dublin, 2020.

[52] Chan V, Estrella MJ, Baddeliyanage R, et al. Rehabilitation among individuals experiencing homelessness and traumatic brain injury: A scoping review. Frontiers in Medicine; 9, https://www.frontiersin.org/articles/10.3389/fmed.2022.916602 (2022, accessed 12 February 2024).

[53] Shoemaker ES, Kendall CE, Mathew C, et al. Establishing need and population priorities to improve the health of homeless and vulnerably housed women, youth, and men: A Delphi consensus study. PLoS One 2020; 15: e0231758.

[54] Betancourt CA, Kitsantas P, Goldberg DG, et al. Substance Use Relapse Among Veterans at Termination of Treatment for Substance Use Disorders. Military Medicine 2022; 187: e1422– e1431.

[55] Padwa H, Bass B, Urada D. Homelessness and publicly funded substance use disorder treatment in California, 2016–2019: Analysis of treatment needs, level of care placement, and outcomes. Journal of Substance Abuse Treatment 2022; 137: 108711.

[56] Health Services Executive Ireland. Alcohol and drug treatment services types. *HSE.ie*, https://www2.hse.ie/living-well/alcohol/coping-difficult-situations/services-types/ (2023, accessed 8 December 2023).

[57] Manning RM, Greenwood RM. Recovery in homelessness: The influence of choice and mastery on physical health, psychiatric symptoms, alcohol and drug use, and community integration. Psychiatr Rehabil J 2019; 42: 147–157.

[58] Moran L, Keenan E, Elmusharaf K. Barriers to progressing through a methadone maintenance treatment programme: perspectives of the clients in the Mid-West of Ireland’s drug and alcohol services. BMC Health Services Research 2018; 18: 911.

[59] Kaur H, Saad A, Magwood O, et al. Understanding the health and housing experiences of refugees and other migrant populations experiencing homelessness or vulnerable housing: a systematic review using GRADE-CERQual. Canadian Medical Association Open Access Journal 2021; 9: E681–E692.

[60] Salem BE, Ma-Pham J. Understanding Health Needs and Perspectives of Middle-Aged and Older Women Experiencing Homelessness. Public Health Nursing 2015; 32: 634–644.

[61] Omerov P, Craftman ÅG, Mattsson E, et al. Homeless persons’ experiences of health-and social care: A systematic integrative review. Health Soc Care Community 2020; 28: 1–11.

